# Identifying undiagnosed high-risk suicidality cases through comorbidity-adjusted risk modeling

**DOI:** 10.1101/2025.07.22.25331824

**Authors:** Louisa Bode, Rena Xu, Matthew Garber, Kenneth D. Mandl, Andrew J. McMurry

**Affiliations:** Computational Health Informatics Program, Boston Children’s Hospital; Peter L. Reichertz Institute for Medical Informatics of TU Braunschweig and Hannover Medical School, Hannover, Germany; Department of Surgery, Harvard Medical School, Boston, MA; Department of Pediatrics, Harvard Medical School, Boston, MA

**Keywords:** Suicide, Public Health Surveillance, Pediatrics, Cohort Selection

## Abstract

**Background:** Suicide is the second leading cause of death for patients aged 10 to 26 years old. Pediatric suicidality is underreported, which poses significant challenges for effective intervention and prevention strategies. Identifying populations at risk for suicidality can provide critical benefits in terms of study cohort selection, prevalence estimation and resource allocation.

**Objective:** (1) Measure prevalence of mental health comorbidities associated with suicidality; (2) propensity match diagnosed suicidality cohorts to select high-risk undiagnosed suicidality cohorts.

**Methods:** ICD-10 diagnosis codes were analyzed for patients aged 6-18 years old presenting to the emergency department at a large academic pediatric hospital between June 1, 2016, and June 1, 2022. Suicidality case definition included subtypes for severity: ideation, self-harm, and attempt. Comorbidities were measured as conditional probabilities of suicidality given a co-occurring ICD-10 diagnosis code. Propensity scores were used to match known suicidality cases to undiagnosed patients at risk of suicidality.

**Results:** In total, 2.9% of ED encounters met an ICD-10-based case definition of suicidality during the study period. Comorbidities of suicidality were statistically significant for 55 frequently co-occurring diagnosis codes. Nearly half (26/55) were not present in the DSM-5 codeset and nearly a quarter (12/55) included ICD-10 codes for harm without documented self-harm intent. The probability of suicidality diagnosis was 44% for patients with personality disorder, gender dysphoria (43%), bipolar disorder (36%), depression (33%), and schizophrenia spectrum disorders (32%). Compared to ground truth comparison, 53.4% of propensity matched comparators were true positive suicidality cases.

**Conclusions:** Propensity score matching is informative for selection of undiagnosed suicidality cases whose comorbidity profiles closely resemble known cases of suicidality.

## Introduction

Suicide is the second leading cause of death among children and adolescents aged 10 to 26 in the United States [1], underscoring a major public health issue. The prevalence of pediatric suicidal ideation, self-harm, and attempts in emergency departments (ED) has dramatically increased [2–4]. However, many at-risk children and adolescents may never receive a formal diagnosis of suicidality, i.e., due to variability in coding practices or stigma-related non-disclosure. Using ICD-10-based case definitions for prevalence estimation or cohort selection may lead to misclassifications or incomplete capture of cases [5–8], ultimately underestimating the true prevalence of suicidal behavior among children and adolescents. Even in cases when the standard Diagnostic and Statistical Manual of Mental Disorders, Fifth Edition [9] is used to guide hospital coding, differences in clinical interpretation and documentation may contribute to undercounting.

Mental health comorbidity plays a crucial role in suicide risk cases [10–12]. Given the link between a higher comorbidity burden and increased suicide rates, it prompts the question [10–12]: Can we utilize information about a patient’s comorbidity burden to identify patients at risk of suicidality that may be systematically undiagnosed in the electronic health record (EHR)? Using a large dataset of ED visits from a major academic pediatric hospital, this study aims to explore the potential of mental health comorbidities as a proxy for identifying underdiagnosed suicidality. The ability to systematically recognize comorbidity-driven risk patterns could enable earlier intervention and more accurate surveillance, resource allocation, and cohort identification for research.

The objectives of this study were (1) measure prevalence of mental health comorbidities associated with pediatric suicidality; and (2) identify high-risk, potential undiagnosed suicidality cohorts using propensity score matching (PSM) and comorbidity risk factors. Source code [13] for reproducing this work is freely available using an open-source Apache 2 License.

## Methods

This retrospective observational study was approved by the institutional review board (IRB-P00043211) at Boston Children’s Hospital. EHR data were extracted and analyzed for the study period June 1, 2016, to June 1, 2022. Selection criteria included all patients aged 6 to 18 years who visited the ED during the study period, identified through the presence of an ED note. Subjects were screened for suicide utilizing the Ask Suicide-Screening Questions toolkit [14]. Patient EHR data were de-identified and loaded into Cumulus [15], a platform for population health studies [16,17].

An ICD-10-based suicidality case definition [5] was used to select patient cohorts who match suicidality criteria. Comorbidities in the suicidality population were reviewed as potential study variables for the ICD-10-based suicidality case definition. Statistically significant associated comorbidities were then used as input variables for a PSM model to select patients who do not match an ICD-10-based suicidality case definition [18]. The propensity matched cohorts were compiled for chart review. Accuracy of the comorbidity-enriched PSM model was then compared to the interpretation of two subject matter experts (RX, AJM).

Figure 1 shows the sequence of case cohort selection, comorbidity analysis, comparator cohort matching, and chart review validation. First, cohorts were selected using an ICD-10-based suicidality case definition. Second, suicidality comorbidities were selected using conditional probabilities, tested for significance, and corrected for multiple hypothesis testing. Third, PSM was used to select patients similar to known suicidality cases. Fourth, manual chart review was used to assess the PSM model identification of suicidality cases missed by a conventional ICD-10-based case selection.

**Figure 1.**
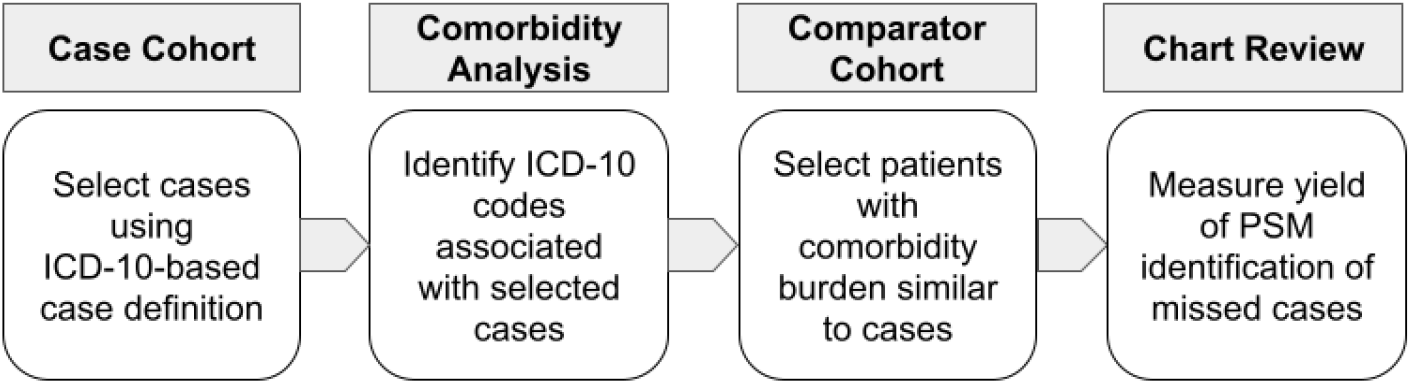
Sequence of case cohort selection, comorbidity analysis, comparator cohort matching, and manual chart review.

### Case Definition

Diagnosis of suicidality included three subtypes with increasing severity: ideation, self-harm, and attempt [6,18]. ICD-10 codesets were compiled for each suicidality subtype [5]. Suicidal ideation is documented only for the present encounter (R45.851), as there are no ICD-10 codes for history of suicidal ideation. Self-harm has four codes for personal history (R45.88, Z91.5, Z91.51, Z91.52) and 1,396 codes in total. Suicide attempt includes four codes: suicide attempt (T14.91), initial encounter (T14.91XA), subsequent encounter (T14.91XD), and sequela of suicide attempt (T14.91XS).

### Study Variables

Study variables include the ICD-10-based suicidality case definition, ICD-10-based diagnosis codes for comorbid DSM-5 disorders, patient demographics, encounter service date, healthcare utilization, and physician note types. Demographic variables include patient reported race, sex assigned at birth, and age at ED visit, with age groups categorized as children (6–11 years) and adolescents (12–18 years). Encounter service date was used to determine the encounter sequence. Healthcare utilization provided a measure of the total number of patient visits and the number of clinical note types per patient. Each encounter was linked to clinical notes including the ED note and optionally psychiatric notes and discharge summary. The statistically significant codes resulting from the comorbidity analysis were added to the list of study variables (see Multimedia Appendix 1).

### Comorbidity Analysis

Conditional probability was used to quantify the relationships between additional ICD-10 diagnoses and the ICD-10-based suicidality case definition. The conditional probability of suicidality “A” given the concurrent presence of a comorbid condition “B” in the same patient (Equation 1) is denoted as

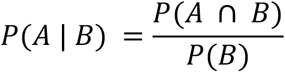

Equation 1. Conditional probability.

A denotes the suicidality ICD-10 codeset. B denotes a particular ICD-10 diagnosis not included in the suicidality codeset.

where P(A ∩ B) signifies the probability of suicidality “A” and diagnosis “B” occurring in the same patient, and P(B) denotes the independent probability of another ICD-10 diagnosis code “B”. Expressed within the range [0, 1], a value of 0 signifies that the diagnosis pairs never co-occur, while a value of 1 indicates that the diagnosis pairs always co-occur.

Conditional probabilities were calculated using Equation 1 for all possible co-occurring ICD-10 diagnosis codes, stratified by age group, sex assigned at birth, and suicidality subtype (ideation, attempt, self-harm). To increase the reproducibility and practical utility of the comorbidity model, only the 20 top-scoring co-occurring ICD-10 codes were selected for each stratification. We then tested the statistical significance of these associations of each stratifier using a Chi-square test for independence, with a Bonferroni correction applied to control for false discovery of multiple comparisons (PsyPy version 1.15.2). ICD-10 codes occurring in fewer than 20 patient cases were omitted from the analysis to reduce the effect of rare events [19]. Conditional probabilities and ICD-10 diagnosis pairs are further referred to as comorbidities even when not all ICD-10 codes necessarily indicate a clinically recognized comorbid condition.

### Propensity Score Matching

PSM was used to match the Case Cohort to a Comparator Cohort covariates across all study variables, i.e., patient demographics (age group, biological sex, race), healthcare utilization codes, clinical note types, DSM-5 categories, and significant ICD-10 codes selected from the comorbidity analysis (Multimedia Appendix 1). The Case Cohort included every patient encounter with a suicidality diagnosis matching the case definition. The Comparator Cohort included every other patient encounter during the study period excluding patients matching the case definition. Case and comparator cohorts were compiled and ordered by encounter date, aggregating study variable observations up to the date of the encounter. To attain larger sample sizes and reduce variation in ICD-10 billing practices, ICD-10 codes were grouped using the Diagnostic and Statistical Manual of Mental Disorders, fifth edition, text revision (DSM-5) [9].

The study variables were compiled into a feature matrix [20] and analyzed using the PsmPy Python package [21]. A logistic regression model estimated propensity scores for suicidality using the feature matrix as input. Similarity between suicidality cases and patients without a suicidality diagnosis was measured using the PSM logit score, i.e., the logarithmic transformation of the predicted probability and k-nearest neighbors.

## Results

### Cohort Characteristics

During the study period, there were 59,866 patients aged 6-18 years old with 90,980 ED encounters in total, of which 2,638 (2.9%) encounters matched an ICD-10-based case definition of suicidality. Table 1 reports demographic characteristics for the overall population and for suicidality cases. Suicidality was significantly more common among females than males across all suicidality subtypes (P <.001 by Chi-square).

**Table 1.** Cohort characteristics.

| Demographics | All ED Encounters (% of Total) | ED Encounters Suicidality Cases (% of Total) | Suicidality Subtypes |  |  |
| --- | --- | --- | --- | --- | --- |
|  |  |  | Ideation | Self-Harm | Attempt |
| Total | 90,980 (100%) | 2638 (100%) | 2275 (100%) | 1030 (100%) | 177 (100%) |
| Age 6-11 y | 47,253 (51.9%) | 310 (11.8%) | 266 (11.7%) | 69 (6.7%) | 5 (2.8%) |
| Age 12-18 y | 43,727 (48.1%) | 2328 (88.2%) | 2009 (88.3%) | 961 (93.3%) | 172 (97.2%) |
| Male | 47,045 (51.7%) | 813 (30.8%) | 707 (31.1%) | 222 (21.6%) | 37 (20.9%) |
| Female | 43,929 (48.3%) | 1825 (69.2%) | 1568 (68.9%) | 808 (78.4%) | 140 (79.1%) |

Figure 2 illustrates the ICD-10-based prevalence of mental health conditions observed in the overall study population according to DSM-5 categories. Anxiety disorders were the most prevalent (15.6%), followed by attention-deficit/hyperactivity disorder (10.4%) and depressive disorders (9.1%). Notably, gender dysphoria (0.6%), dissociative (0.4%), and personality disorders (0.3%) were rare DSM-5 categories.

**Figure 2.**
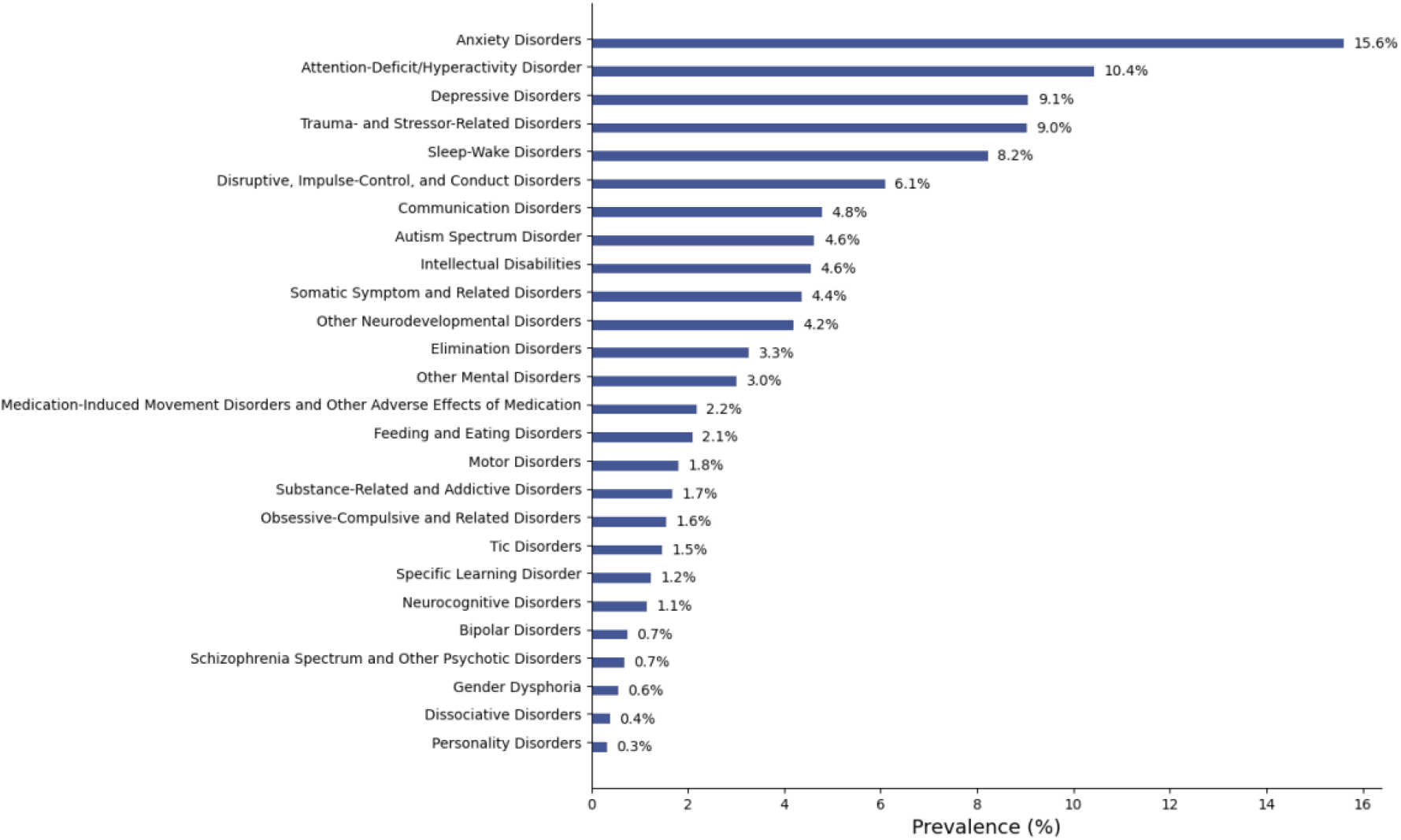
Prevalence of DSM-5-categorized mental health conditions. Y-axis denotes each DSM-5 category. X-axis denotes the ICD-10 code-based prevalence of the DSM-5 categorized mental health disorders. The DSM-5 categories “paraphilic disorders” and “sexual dysfunctions” are not displayed due to the low prevalence (≤0.1%).

### Comorbidity Analysis

Comorbidity analysis revealed the conditional probability of suicidality was highest for these DSM-5 categories: personality (44.2%), gender dysphoria (42.9%), bipolar (36.1%), depression (33.9%), and schizophrenia spectrum (32.4%). Conversely, the probability of DSM-5 category given suicidality was highest for depression (82.6%), anxiety (71.6%), trauma and stressor (36.0%), attention deficit disorder (ADD) and attention deficit hyperactivity disorder (ADHD) (32.3%), and disruptive impulse control and conduct (27.3%). Roughly half (50.5%) of all patients with an ICD-10 code for gender identity also met the suicidality case definition. Figure 3 illustrates the conditional probabilities of suicidality and DSM-5-categorized mental health conditions. Detailed statistics for each DSM-5 category are found in Multimedia Appendix 2.

**Figure 3.**
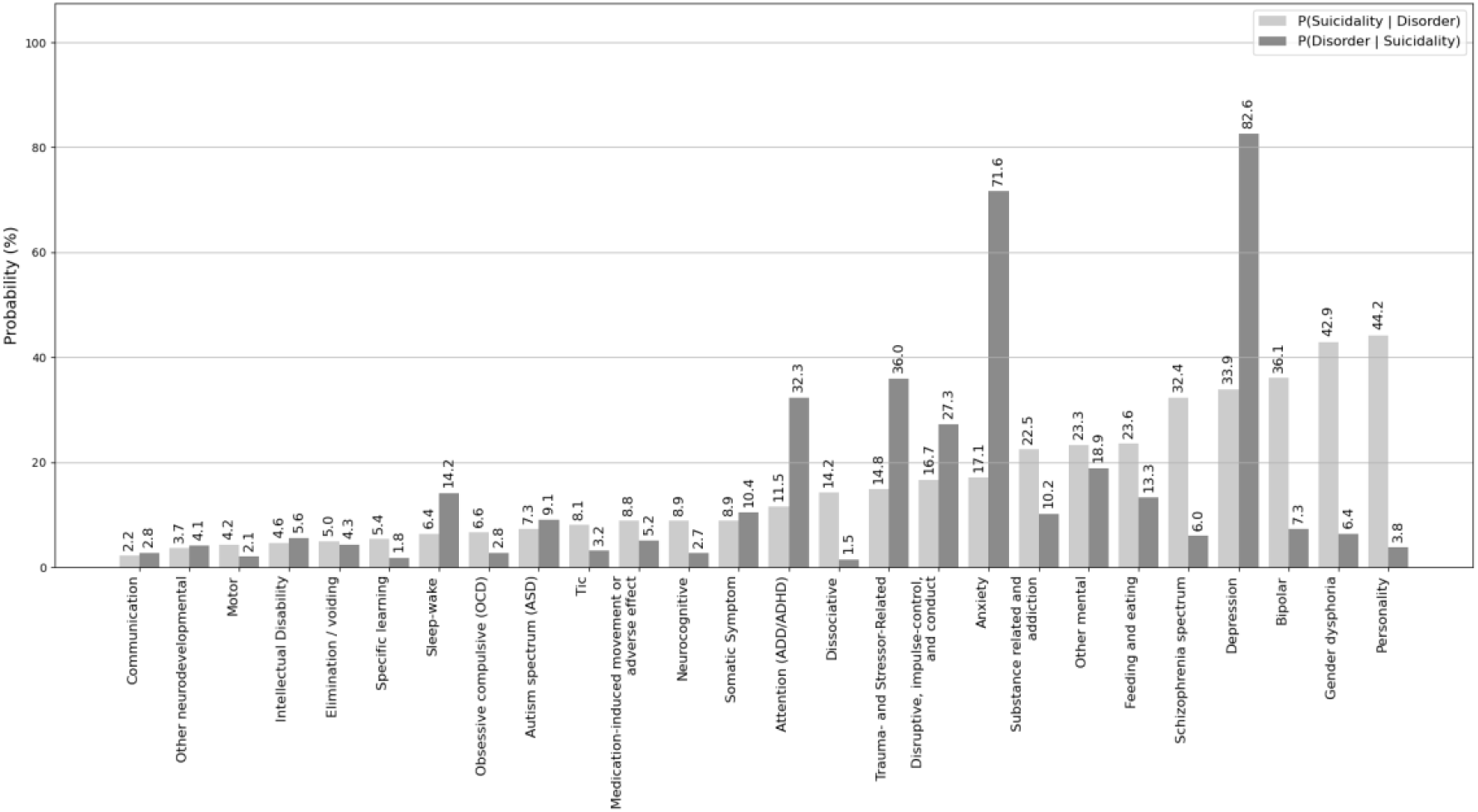
Conditional probabilities of suicidality and DSM-5-categorized mental health conditions. The light gray bars show the probability of suicidality given a specific disorder, while the dark gray bars show the probability of a disorder given that individuals also have a documented ICD-10 code for suicidality.

Of the 55 statistically significant ICD-10 suicidality risk factors, nearly half (26/55) were not part of the DSM-5 standard mental health diagnosis manual (Multimedia Appendix 1). Nearly a quarter (12/55) were for harm without documented self-harm intent, including “unintentional” drug poisoning and “accidental” lacerations of arms and legs. These ICD-10 codes for unintentional poisoning or accidental injury were strongly associated with self-harm diagnosis. Female patients were significantly more likely (OR of 2.23 (95% CI: 1.61–3.10, P <.001)) than male patients to have ICD-10 codes related to accidental (unintentional) or undetermined self-harm. For a more detailed report on findings between subpopulations, see Multimedia Appendix 3.

Mental health conditions such as F48.9 (nonpsychotic mental disorder, unspecified), F43.12 (post-traumatic stress disorder, chronic), and R44.0 (auditory hallucinations) were not found in DSM-5 classifications [9] yet were significant suicidality risk factors within the study population. About half (50.5%) of all patients with an ICD-10 code for gender identity (F64.8) also had an ICD-10 code for suicidality. A full list of the top-scoring risk factors of suicidality are shown in Multimedia Appendix 4.

### Propensity Score Matching

PSM was used to match cases with comparators sharing similar comorbidity burdens that may underreport cases using an ICD-10-based case selection. Figure 4 illustrates the distribution of propensity scores for suicidality cases and comparators before and after matching. Propensity scores closer to 0 indicate lower likelihood of suicidality, while scores closer to 1 represent higher likelihood. Before matching, 70.4% of suicidality cases (Fig. 4a) had a propensity score ≥0.9. By contrast, only 1.85% of comparators had scores ≥0.9 (Fig. 4b). After matching, the distribution of matched comparators (Fig. 4d) more closely resembled that of the cases. The number of comparators with propensity scores <0.5 dropped from 94.7% before matching to 12.2% after matching, demonstrating that PSM was effective in aligning the comparator group’s comorbidity profile with that of the suicidality cases. For more details on the PSM analysis see Multimedia Appendix 5.

**Figure 4.**
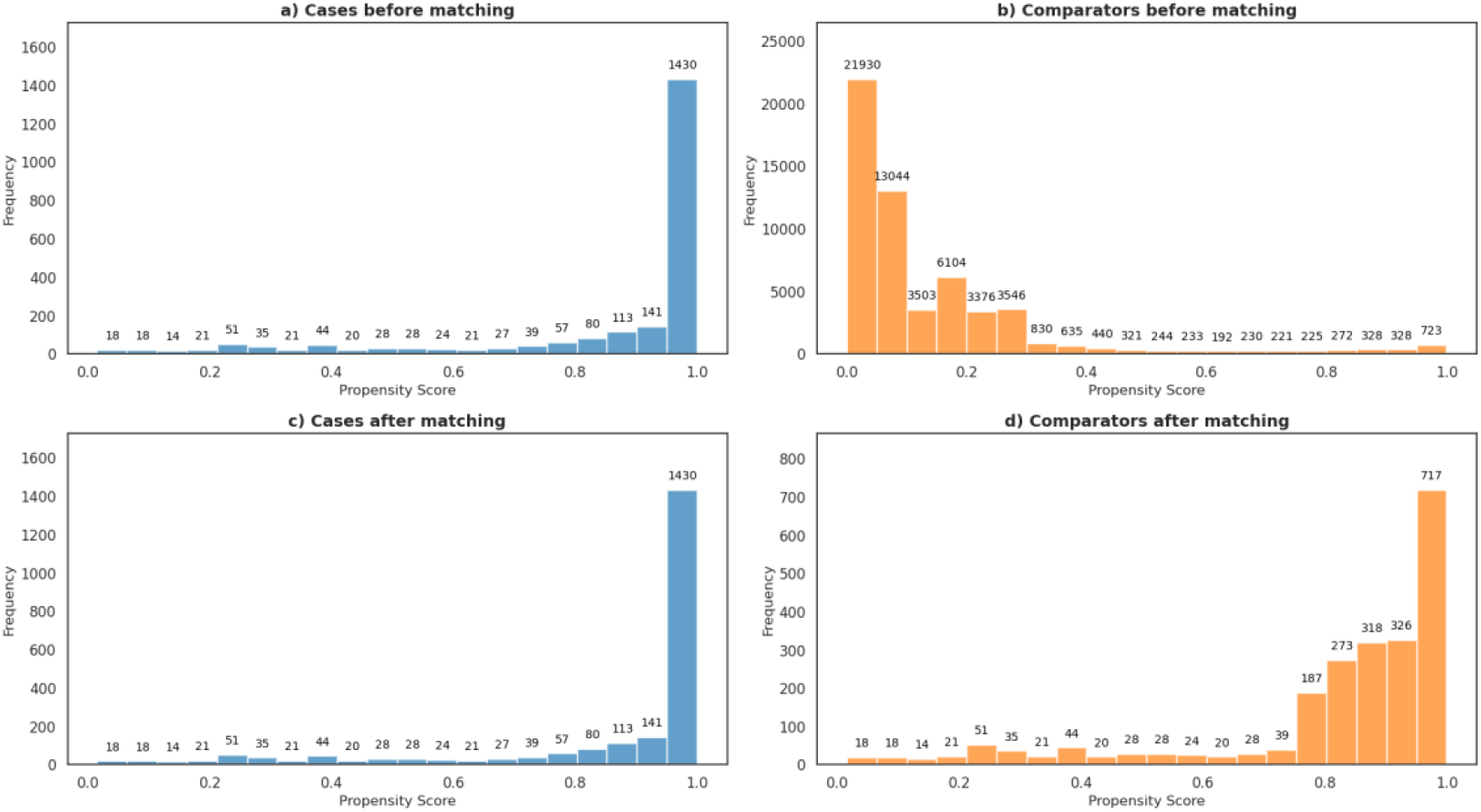
Distribution of propensity scores for suicidality cases and comparators before and after matching. X-axis denotes propensity scores. Y-axis denotes the frequency representing the size of the patient population.

Propensity matched cohorts were selected for chart review (N=205 encounters) by subject matter experts [5]. Compared to the prevalence of suicidality in the overall study population (2.9%), the prevalence in the PSM-matched cohort was 22.3%. The difference of proportions of suicidality in the two populations was significant (P<.001 two proportions z-test).

## Discussion

In this study, a patient matching approach using comorbidity profiles identified a population of patients with high prevalence of suicidality despite lacking ICD-10 codes for suicidality. This finding has several important implications. First, the propensity score matching methodology could enable more accurate cohort construction for suicidality research, public health surveillance, and population-level analyses of suicidality risk, prevalence, and outcomes. ICD-10 codes alone may not be reliable for identifying cases of suicidality [5]. By demonstrating the utility of PSM to identify potential underdiagnosis and/or inadequate documentation of suicidality, this study advances methods for improving case ascertainment.

These findings have important clinical implications as well. This study identified comorbidity patterns that are associated with increased probability of suicidality. In patients with these phenotypes, additional screening may be warranted to assess for suicide risk. For instance, patients with multiple ED encounters for supposedly unintentional self-harm may be at increased risk for suicidality and require more in-depth psychiatric evaluation than the standard suicide screening measures employed for all ED patients. Moreover, while the objective of this study was to develop a method to identify “unrecognized” cases of existing suicidality rather than to predict future risk of suicidality, it is feasible that the high-risk comorbidity patterns identified in this study are also associated with increased risk of future suicidality. Additional research is needed to understand how different comorbidity patterns predict suicidality risk and how such insights could be operationalized to improve care and outcomes.

Public health surveillance of pediatric suicidality using ICD-10 codes is likely underreported. In the chart reviewed sample, only about half (53.4%) of suicidality cases were identified using ICD-10 codes. While it was beyond the scope of this study to estimate the true and apparent prevalence [22] of suicidality, it may be possible to use large language models to provide a more accurate prevalence estimation [23].

There were limitations in this retrospective observational study using EHR data. First, this study was conducted in a single large pediatric medical center in a northeastern urban area. Clinically relevant comorbidity profiles may vary by geography and urban versus rural settings. Second, this study focused on suicidality in ED encounters. ICD-10 coding practices may vary across ED, inpatient, and ambulatory settings. Third, while propensity score matching identified many instances of suicidality that were missed using ICD-10 codes alone, it still might not identify all suicidality cases. Fourth, the effectiveness of propensity score matching is highly dependent on the selected covariates [24]. Unmeasured or uncoded diagnoses could not be accounted for, potentially limiting the accuracy of matched comparisons. To maintain clinical interpretability and feasibility, we restricted the model to top-ranking comorbidities that a clinician could reasonably monitor in practice. However, other significant comorbidities not included in the model may also influence suicidality risk and affect matching accuracy. Further studies to test and refine the methodology may help to increase the case detection yield.

## Conclusions

This study provides a method for selecting case matching cohorts with pediatric suicidality. Variables for the matched selection include significantly associated suicidality comorbidities and demographic variables in the study population. These findings suggest that pediatric suicidality may be highly underreported using an ICD-10-based case definition of suicidality.

## Supporting information

Appendix 1: Significant suicidality risk factors

Appendix 2: DSM-5 Disorder Prevalence

Appendix 3: Findings of suicidality subpopulations

Appendix 4: Top 20 ICD-10 codes by sex, age, and subtype

Appendix 5: Propensity Score Matching

## Data Availability

All data produced in the present work are contained in the manuscript

https://github.com/smart-on-fhir/cumulus-library-suicidality-icd10

## Acknowledgements

AM, RX, and KM contributed to the study design. AM and RX conducted the chart review. LB and AM drafted the manuscript, with RX, KM, and MG reviewing and editing the manuscript. LB performed the data analysis, and MG implemented the open-source code.

We thank our colleagues Tim Miller and Karen Olson for their advice on the study methodology. We also thank Alon Geva for his support with the chart review.

This study was supported by the Centers for Disease Control and Prevention (CDC) of the U.S. Department of Health and Human Services (HHS) as part of a financial assistance award. The contents are those of the authors and do not represent the official views of, or an endorsement by, CDC, HHS, or the U.S. government. Support was provided by the National Center for Advancing Translational Sciences, National Institutes of Health Cooperative Agreement (U01TR002623).

## Abbreviations

DSM-5: Diagnostic and Statistical Manual of Mental Disorders 5th Edition
EHR: electronic health record
ED: emergency department
ICD-10: International Classification of Diseases 10th Revision
PSM: Propensity Score Matching

## Multimedia Appendix

Multimedia Appendix 1: Significant suicidality risk factors

Multimedia Appendix 2: DSM-5 Disorder Prevalence

Multimedia Appendix 3: Findings of suicidality subpopulations

Multimedia Appendix 4: Top 20 ICD-10 codes by sex, age, and subtype

Multimedia Appendix 5: Propensity Score Matching

