## Appendix 3: Findings of suicidality subpopulations for "Identifying undiagnosed high-risk suicidality cases through comorbidity-adjusted risk modeling"

### Suicidality Subpopulation Findings

The methodologies designed in this study address the lack of a validated suicidality case definition and the potential underreporting of suicidality cases from a public health perspective. In addition, it expands upon existing research by overcoming current challenges with ICD-10 and minimizing efforts in manual annotation. PSM was used to propose potential cases to human expert reviewers to estimate how much the potential undercounting is. A sensitivity below 58% suggests that suicidality prevalence may be vastly underreported. Validated by chart review, the evaluation metrics indicate that while ICD-10 codes are useful in identifying suicidality, they should be supplemented with other cohort selection methods or, when feasible, clinical judgment. PSM can help refine an ICD-10-based EHR search by identifying a substantial proportion of additional cases, thus increasing the sensitivity of detecting a wider range of suicidality manifestations. In summary, the chart review results reveal a considerable gap in detecting true cases of suicidality, however, these metrics may vary based on the population under study, the specific search criteria or suicidality definitions applied, and other factors including the expertise of the reviewers.

The findings can be informative to clinicians, as they identify comorbidities most frequently co-occurring with cases of suicidality, pinpointing risk factors and highlighting ICD-10 coding practices. From those frequently co-occurring comorbidities that were identified, the majority

of ICD-10 codes refer to the heading “mental, behavioral and neurodevelopmental disorders”

(F01-F99). Other notable codes include "symptoms, signs and abnormal clinical and laboratory findings" (R00-R99), "factors influencing health status and contact with health services" (Z00-Z99), and "injury, poisoning and certain other consequences of external causes" (S00-T88).

Among all self-harm patients, accidental poisoning (T43.201A, T39.1X4A, T65.94XA) and injuries (S50.812A, S61.512A) ranked highest. While these codes may not inherently denote an intentional act of self-harm, they serve as valuable indicators to identify patients demonstrating self-harming tendencies, extending beyond those explicitly recognized for intentional self-harm.

The ICD-10 codes most associated with suicidality among both males and females include unspecified nonpsychotic mental disorder (F48.9), severe major depressive episode without psychotic features (F32.2, F33.2), homicidal ideations (R45.850), and problems related to lifestyle (Z72.89). In the female suicidality population, there is a link to ICD-10 codes that represent "accidental" or "undetermined" poisonings by some toxic substance (e.g., T43.201A and T39.1X4A). Codes related to external causes (e.g., poisoning, superficial injuries, or open wounds of specific body parts (S60-S79)) are part of the case definition but only if they are specified as "intentional" self-harm. Nonetheless, these self-harm codes rank higher in conditional probability among females than males, as well as among adolescents than children, regardless of whether they reflect coding errors. In the male population, suicidality is particularly an issue among those who identify differently from their assigned sex at birth, indicated by codes for transsexualism (F64.0) and unspecified gender identity disorder (F64.9).

Conditional probabilities observed in patients with one or more suicide attempts are relatively lower when compared to other suicidality subtypes. This may be attributed to the overall low prevalence of suicide attempts in the broader population. Comorbidities including chronic post-traumatic stress disorder (F43.12), eating disorders (F50.9), and anxiety disorders (F41.1, F41.8) often co-occur with suicide attempts.

ICD-10 chapter “mental and behavioural disorders”, including F48.9, F33.2, and F32.2 most often co-occur with suicidal ideation. Low self-esteem (R45.81), bipolar II disorder (F31.81), and homicidal ideations (R45.850) are among the top 20 comorbidities identified in patients with suicidal ideation, distinguishing these individuals from those who have attempted suicide or engaged in self-harm. In addition, a strong and persistent cross-gender identification coupled with persistent discomfort with their sex (F64.8, F64.9) is with almost 45% a risk factor towards suicidal ideation.
