## Appendix 5: Propensity Score Matching for "Identifying undiagnosed high-risk suicidality cases through comorbidity-adjusted risk modeling"

Propensity score matching (PSM) is a method for balancing observed covariates in observational studies [1], ensuring that the post-matching distributions of these covariates between treated and untreated subjects are analogous, thus mitigating confounding effects. Defined as the conditional probability of being assigned to a particular treatment given observed baseline covariates [1], the propensity score is used to assemble pairs comprising treated and untreated subjects based on their similarity in propensity scores. In this study, PSM was chosen as the method to select suicidality cohorts by matching patients lacking an ICD-10 code for suicidality and who share similar comorbidity profiles with those who do meet the case criteria of suicidality, potentially increasing ascertainment of suicidality cases in public health studies.

The effectiveness of the PSM model in reducing the effect sizes of each covariate among patients who have a recorded ICD-10 code for suicidality and those who do not is illustrated in [2]. Effect size plots, utilizing effect size metrics such as Cohen’s d, show the magnitude of observed effects. However, a decrease in effect sizes for all covariates is not always guaranteed, e.g., in the case of "Intellectual Disabilities", where the effect size increased post-matching [2]. Patients with intellectual disabilities were more disproportionately represented in one of the two groups even after the matching process, as the limited number of patients with these conditions may pose challenges in simultaneously balancing all other covariates during the matching process. Low-prevalence disorders can make it more challenging to achieve complete balance across all covariates in the PSM model. Achieving a perfect balance between the two groups is not the primary objective of this study. The primary reason to use PSM here is rather to select patients who might have a missed diagnosis for suicidality and measure the reliability of ICD-10 suicidality codes for population health studies. Minor imbalances in covariates can be deemed uninformative and do not significantly impact the overarching objectives of the study.

After training the logistic regression model, a probability referred to as the Propensity Score (PS), was computed for each subject using the included covariates. Figure 4 illustrates the distributions of PS for patients with (cases) and without a documented ICD-10 code for suicidality (comparators) before and after matching. Matched comparators with PS close to 1.0 are more likely to be false negatives, useful in understanding both sensitivity and case selection with regards to missed cases, whereas known cases with PS close to 0 are more likely false positives, negatively impacting the PPV. Subjects with similar or identical PS from either group are paired to minimize disparities in observed characteristics between the two groups. The matching algorithm systematically seeks comparators with analogous PS for all cases. Adjusting the caliper distance allows for controlling how large the difference between propensity scores can be when matching cases and controls, thereby selecting fewer or greater numbers of potential matches. When using matching without replacement, there is a lack of comparable counterparts with a similar PS. In simpler terms, if an exact match for every case cannot be found, the distribution after matching may not precisely mirror the distribution before matching. In this study, 717 comparators with a PS close to 1 were available, however, were insufficient to match the 1430 cases with an identical or similar PS close to 1 (see Figure 4c and 4d).
